# Estimating the seroincidence of scrub typhus using antibody dynamics following infection

**DOI:** 10.1101/2022.11.07.22282017

**Authors:** Kristen Aiemjoy, Nishan Katuwal, Krista Vaidya, Sony Shrestha, Melina Thapa, Peter Teunis, Isaac I. Bogoch, Paul Trowbridge, Pacharee Kantipong, Stuart D. Blacksell, Tri Wangrangsimakul, George M Varghese, Richard Maude, Dipesh Tamrakar, Jason R. Andrews

**Affiliations:** Department of Public Health Sciences, University of California Davis School of Medicine, Davis, CA, USA; Department of Microbiology and Immunology, Faculty of Tropical Medicine, Mahidol University, Bangkok Thailand; Dhulikhel Hospital, Kathmandu University Hospital, Dhulikhel, Nepal; Center for Global Safe Water, Sanitation and Hygiene, Hubert Department of Global Health, Rollins School of Public Health, Emory University, Atlanta, GA, USA; Department of Medicine, University of Toronto, Toronto, ON, Canada; Michigan State University School of Human Medicine, Grand Rapids, MI, USA; Department of Internal Medicine, Chiangrai Prachanukroh Hospital, Chiang Rai, Thailand; Mahidol-Oxford Tropical Medicine Research Unit, Faculty of Tropical Medicine, Mahidol University, Bangkok Thailand; Centre for Tropical Medicine and Global Health, Nuffield Department of Medicine, University of Oxford, Oxford, UK; Department of Infectious Diseases, Christian Medical College, Vellore, India; The Open University, Milton Keynes, UK; School of Public Health, Li Ka Shing Faculty of Medicine, University of Hong Kong, Hong Kong Department of Medicine, Stanford University School of Medicine, Stanford, CA, USA; Division of Infectious Diseases and Geographic Medicine, Stanford University School of Medicine, Stanford, CA, USA

**Keywords:** Orientia tsutsugamushi, Scrub Typhus, Antibody kinetics, Seroepidemiology, incidence

## Abstract

**Introduction:** Scrub typhus is an acute febrile illness caused by the bacterium *Orientia tsutsugamushi*. Characterizing the population-level burden of scrub typhus is challenging due to the lack of accessible and accurate diagnostics. In this study, we present a novel approach that utilizes information on antibody dynamics after infection to generate population-level scrub typhus seroincidence estimates from cross-sectional serosurveys.

**Methods:** We use data from three cohorts of scrub typhus patients enrolled in Chiang Rai, Thailand, and Vellore, India, and representative population data from two serosurveys in and around the Kathmandu valley, Nepal, and Vellore, India. The samples were tested for IgM and IgG responses to *Orientia tsutsugamushi*-derived recombinant 56-kDa antigen using commercial ELISA kits. We used Bayesian hierarchical models to fit two-phase models to the antibody responses from scrub typhus cases and used the joint distributions of the peak antibody titers and decay rates to estimate population-level incidence rates in the cross-sectional serosurveys. We compared this new method to a traditional cut-off-based approach for estimating seroincidence.

**Results:** Median IgG antibodies persisted above OD 1.7 for 22 months, while IgM displayed longer persistence than expected, with 50% of participants having an OD >1 for 5 months. We estimated an overall seroincidence of 18 per 1000 person-years (95% CI: 16-21) in India and 4 per 1000 person-years (95% CI: 3-6) in Nepal. Among 18 to 29-year-olds, the seroincidence was 8 per 1000 person-years (95% CI 4 -16) in India and 9 per 1000 person-years (95% CI: 6-14) in Nepal. In both India and Nepal, seroincidence was higher in urban and periurban settings compared to rural areas. Compared to our method, seroincidence estimates derived from age-dependent IgG-seroprevalence without accounting for antibody decay underestimated the disease burden by 50%. By incorporating antibody dynamics, the approach described here provides more accurate age-specific infection risk estimates, emphasizing the importance of considering both IgG and IgM decay patterns in scrub typhus seroepidemiology.

**Conclusion:** The sero-surveillance approach developed in this study efficiently generates population-level scrub typhus seroincidence estimates from cross-sectional serosurveys. This methodology offers a valuable new tool for informing targeted prevention and control strategies, ultimately contributing to a more effective response to scrub typhus in endemic regions worldwide.

## Introduction

Scrub typhus, an acute febrile illness caused by the bacterium *Orientia tsutsugamushi (OT)*, is an important, under-recognized etiology of fever (1). Once thought to be restricted to the “tsutsugamushi triangle”, a region spanning from Russia to Pakistan, Australia, and Japan, recent studies have identified scrub typhus transmission in South America, Africa, and the Middle East (2–4). Infections occur when trombiculid mite chiggers (larvae) enter a host’s skin through hair follicles and feed on lysed skin tissue. The mites, both vectors and reservoirs of OT, feed on various mammals, including humans and rodents (5). In humans, symptoms are non-specific and include fever, myalgia, headache, gastrointestinal symptoms, and rash. An eschar is found at the inoculation site in a variable proportion of cases and is frequently missed on clinical examination. Case fatality rates are estimated to be 1-2% among treated patients and 6% among untreated patients (6).

Determining where scrub typhus transmission occurs is critical to inform public health interventions and research priorities. Clinical incidence underestimates the true underlying burden of disease due to non-specific symptoms and the lack of accessible and accurate diagnostics (7). Periodic serosurveillance studies across endemic countries have demonstrated significant heterogeneity in seroprevalence within and between countries (8). However, directly comparing seroprevalence is not straightforward because of differences in the age distributions of each sampled population and uncertainty in antibody-waning patterns.

Here, we apply a novel analytic approach to estimate scrub typhus seroincidence using antibody decay information from confirmed cases. The decay of antibody concentrations defines a timescale for inferring when an infection occurred. This approach does not depend on classification using cutoffs, which have been difficult to derive for scrub typhus across locations with varying forces of infection (9). We first model longitudinal IgG and IgM antibody responses to OT-derived antigens among confirmed scrub typhus cases in Thailand and India and then use these parameters to estimate scrub typhus seroincidence from cross-sectional population serosurveys in Nepal and India.

## Methods

### Study populations and enrollment

#### Scrub typhus cases

We used antibody responses measured from confirmed scrub typhus patients enrolled in three studies: two in Thailand and one in India. The first study in Thailand enrolled children less than 18 years old who were admitted to Chiang Rai Prachanukroh Hospital with fever (temperature higher than 37.5°C) or a history of fever within the last 14 days(10). Enrollment occurred between July 2015 to August 2016. Confirmed scrub typhus infections were defined by meeting at least one of the following criteria: (1) a positive PCR or culture result from either a blood or eschar sample, (2) a fourfold rise in the IgM titer to ≥1:3200 in paired serum or plasma samples, or (3) a single IgM titer of ≥1:3200 in an acute-phase serum or plasma sample(11,12). Blood samples were collected at baseline and at 2-, 12-, and 52-weeks after enrollment, stored at −80°C, and transported to Bangkok for diagnostic processing.

The second study In Thailand enrolled hospitalized patients older than 15 years from a fever surveillance study also at the Chiang Rai Prachanukhao Hospital (13,14). Enrollment occurred from August 2007 to August 2008. Patients confirmed to be infected with scrub typhus were defined by meeting at least one of the following criteria: (1) *In vitro* isolation of *O. tsutsugamushi*, (2) a ≥4-fold rise in IgM titer in paired serum samples when tested by the indirect immunofluorescence assay, (3) a positive result in at least two out of the three PCR assays described in Paris et al., 2011(14). Serum samples were collected at admission and stored at −80°C until testing.

In India, serum samples were collected from individuals infected with scrub typhus who were > 18 years old and sought care at Christian Medical College Teaching Hospital in Vellore, India, between December 2011 and March 2015 (15). Confirmed patients with scrub typhus were defined by a positive IgM ELISA (optical density (OD) >0.8) and/or a positive PCR for *O. tsutsugamushi*.

The serum samples were collected cross-sectionally from retrospective cases with diagnoses between 2 months and 3.5 years prior. Serum samples were collected in the patient’s household, stored at ^−^80^0^C, and processed within two weeks of collection.

#### Population samples

In Nepal, we enrolled a geographically random, population-based cross-sectional sample of individuals aged 0 to 25 years from the catchment areas of Kathmandu University Teaching Hospital in Kathmandu (urban), and Dhulikhel Hospital in Kavrepalanchok, Nepal (periurban and rural) (16). Within catchment areas, we randomly selected geographically defined grid clusters and enumerated all households in each cluster. From this census, we randomly selected individuals and sought consent. We collected capillary blood samples from consenting participants onto TropBioTM filter papers (Cellabs Pty Ltd., Brookvale, New South Wales, Australia). The samples were air-dried for at least two hours at room temperature, then stored with desiccant in individual plastic bags at -20 °C until processing. Study participants were enrolled between February 2019 and Jan 2021. In India, we utilized a previously-conducted cross-sectional serosurvey for scrub typhus conducted from September 2014 to December 2014(17).

The study enrolled adults ≥18 years of age in the Vellore District of Tamil Nadu in South India. A two-stage clustered sample design was used to randomly select communities and individuals(17).

Venous blood was collected from consenting participants, transported on ice, and stored at -70°C until tested.

### Laboratory methods

All samples were tested using IgM and IgG responses to *O. tsutsugamushi*-derived recombinant 56-kDa antigen using the Scrub Typhus Detect ELISA kit (InBios International, Inc., Seattle, WA, USA)(13) performed as per the manufacturer’s instructions. All serum samples were tested at a 1:100 dilution, and the results were read at 450 nm using a microplate reader (Thermo Scientific Multiskan FC) to generate a final optical density result (optical density (OD) at 450 nm). To prepare the dried capillary blood samples used in Nepal, we cut two filter paper protrusions and submerged them in 133 *μ*L of 1XPBS 0.05% Tween buffer overnight at 4 C, then centrifuged to recover the eluates. The eluate was assumed to be equivalent to a 1:10 dilution of plasma.

### Statistical methods

We estimated seroincidence in two ways: 1) by deriving it from the age-dependent seroprevalence and 2) as a function of the longitudinal antibody dynamics after infection. For method 1, we used finite mixture models to determine the IgG seropositivity cutoffs using the mean plus two standard deviations of the first mixture component (18). We calculated IgG seroprevalence as the proportion of individuals classified as IgG seropositive. We then used an exponential survival model to derive seroincidence (equations 1 & 2), where *π*(*a*) equals the seroprevalence at age (*a*) and *λ* equals the seroincidence rate.

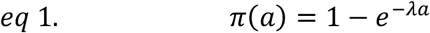

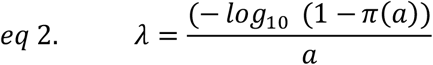

We fit a generalized linear model to the binomial seropositivity outcome conditional on age with a complementary log-log link and estimated seroincidence from the model’s intercept term (19,20). This approach assumes there is no antibody waning over time and that seroincidence is constant over age. To evaluate how seropositivity changes over age, we fit generalized additive models (21) with a cubic spline for age and simultaneous confidence intervals using a parametric bootstrap of the variance-covariance matrix of the fitted model parameters (22).

For method 2, we used information about antibody decay from confirmed cases to estimate seroincidence. First, we modeled longitudinal antibody dynamics after scrub typhus infection using two-phase models with an infection episode characterized by an exponential rise in antibody response and a non-exponential power function decay (23–25). We model antibody kinetics from onset of fever (*t* = 0) with initial antibody levels of *y*_0_ reaching a peak antibody response of *y*_1_. We used a Bayesian hierarchical framework to fit the above models for IgG and IgM (23,24) and generate joint distributions using Markov chain Monte Carlo (MCMC) sampling, allowing for individual variation. We implemented the models in R version 4.1.3 using JAGS(26).

We used the distributions of peak antibody response and the decay rate and shape to estimate seroincidence in the cross-sectional population samples(27). We created a likelihood function for the observed cross-sectional population data based on the longitudinal kinetics following scrub typhus infection with the assumption that incident infections occur as a Poisson process with rate lambda (*λ*(28). We generated maximum likelihood profiles for *λ* using each isotype separately and jointly by combining their likelihood functions. We accounted for two sources of noise in the observed serologic responses: measurement noise of the assay and biologic noise as detailed in Teunis et. al. (27).

### Ethics statement

Institutional Review Boards in India (Christian Medical College, Vellore), Thailand (Chiang Rai Hospital, the Faculty of Tropical Medicine, Mahidol University, and the Thai Ministry of Public Health), The United States (Stanford University Institutional Review Board), and Nepal (Nepal Health Research Council Ethical Review Board)) approved the study forms and protocols.

## Results

ELISA antibody responses were measured from 288 patients with confirmed scrub typhus infections: 77 in Thailand and 211 in India (15). Samples were collected between 0 and 1300 days after symptom onset. In the pediatric cohort in Thailand, 4 samples were collected between 0 400 days after symptom onset. In the adult cohorts in India and Thailand, just one sample was collected between 1 and 11 days after symptom onset (median 5.5) in Thailand and between 65 and 1250 days after symptom onset (median 599) in India. The adult cohort in Thailand did not measure IgG responses. The median ages of participants were 44 years (IQR 27-55, range 1.2 - 84); 47 in India (IQR 35-57), and 43 in the adult cohort in Thailand (IQR 35-51) and 6.6 (IQR 4 – 10) in the pediatric cohort in Thailand (Table 1).

**Table 1:** Summary of longitudinal scrub typhus patient data.

|  | India: Adult<br>(N=211) | Thailand:<br>Pediatric<br>(N=35) | Thailand:<br>Adult<br>(N=42) | Overall<br>(N=288) |
| --- | --- | --- | --- | --- |
| **Age (years)** |  |  |  |  |
| Median (IQR) | 47 ( 35 - 57) | 6.6 ( 4.0 - 10) | 43 (35 - 51) | 44 (27 - 55) |
| Min/Max | 19 - 79 | 1.2 - 13 | 15 - 84 | 1.2 - 84 |
| Missing | 0 (0%) | 0 (0%) | 1 (2.4%) | 1 (0.3%) |
| **Days from symptom onset to first visit** |  |  |  |  |
| Median (IQR) | 520 (340 - 880) | 10 (7.5 - 12) | 5.5 (4.0 - 7.0) | 380 (13 - 750) |
| Min/Max | 65 - 1300 | 3.0 - 19 | 0.15 - 11 | 0.15 - 1300 |
| **Days from symptom onset to last visit** |  |  |  |  |
| Median (IQR) | - | 350 ( 350 - 370) | - | 350 ( 350 - 370) |
| Min/Max |  | 10 - 400 |  | 10 - 400 |
| **Number of visits** |  |  |  |  |
| Median (IQR) | 1.0 ( 1.0 - 1.0) | 4.0 ( 4.0 - 4.0) | 1.0 ( 1.0 - 1.0) | 1.0 ( 1.0 - 1.0) |
| Min/Max | 1.0 - 1.0 | 1.0 - 4.0 | 1.0 - 1.0 | 1.0 - 4.0 |

Median IgG responses were elevated above the mixture model cut-off for 22.3 months after symptom onset (Figure 2A). An OD of 1 has been suggested as a reasonable cutoff to indicate a recent infection (29,30). In our study population, median responses were elevated about an OD of 1 for 5.3 months after infection, 25% of the population had elevated for 12.5 months and 10% had elevated responses for 26.5 months. IgM responses decayed at a rate of OD 1.7 (95% credible interval (CrI) 0.77 – 3.77) per month compared to 0.11 (95% CrI 0.04 – 0.29) per month for IgG). IgM responses peaked at OD 5.12 (95% CrI 4.00 – 7.03), 5.2 days (95% CrI 3.4 – 7.6) after symptom onset. IgG responses peaked at OD 2.8 (95% CrI 2.1 – 3.73) 11 days (95% CrI 5.7 – 20.6) after symptom onset. (Supplemental Table 2).

**Figure 1:**
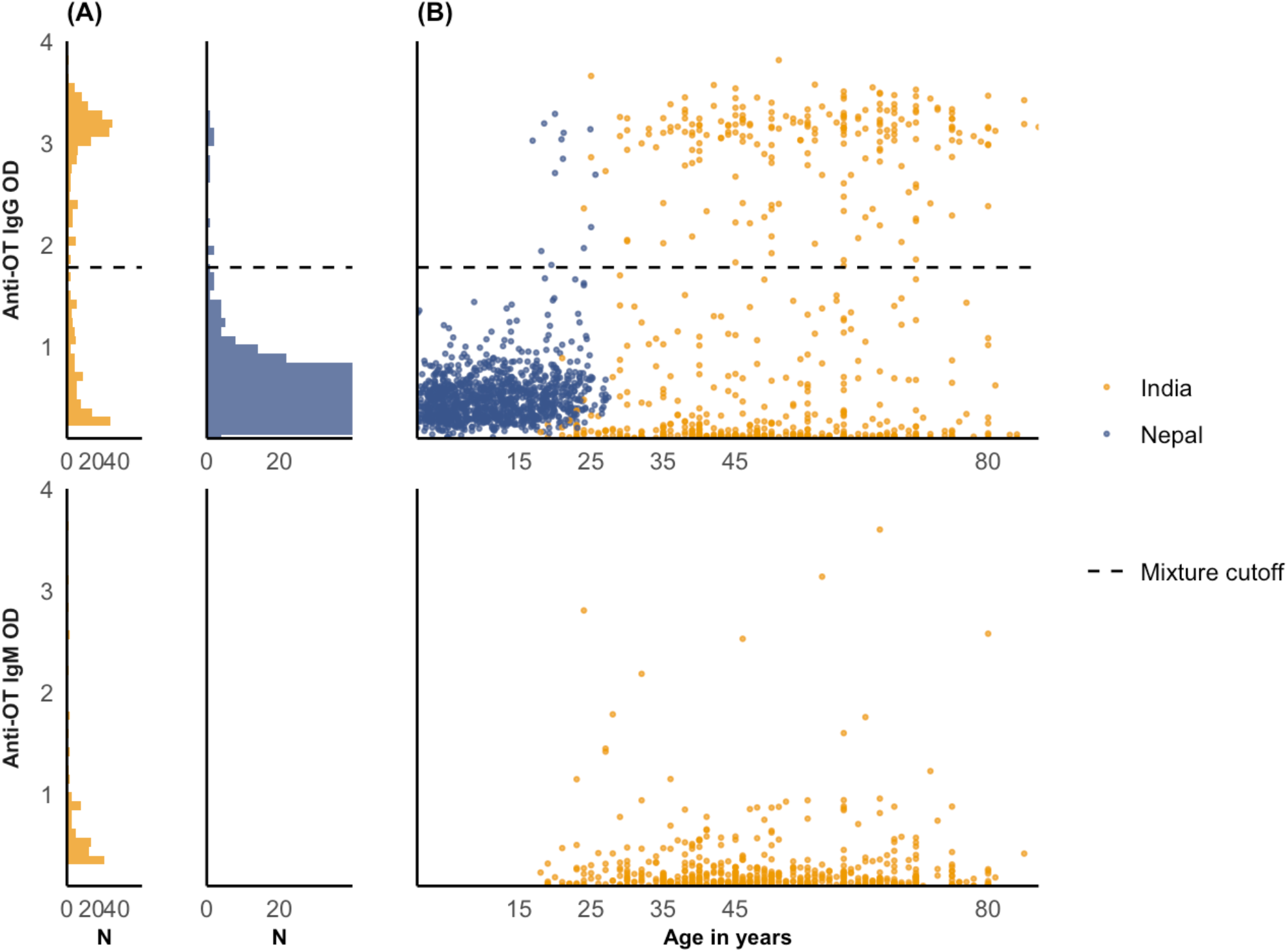
Quantitative antibody responses among population serosurveys in Nepal and India. Anti-OT IgM and IgG responses are measured using kinetic enzyme-linked immunosorbent assays (ELISAs) and depicted on the y-axis of all plots. The horizontal dashed line denotes the mixture model cutoff. Panel A shows the distribution of responses among the population-based samples in India and Nepal. Panel B shows the antibody among the population-based samples in India and Nepal as a function of age.

**Figure 2:**
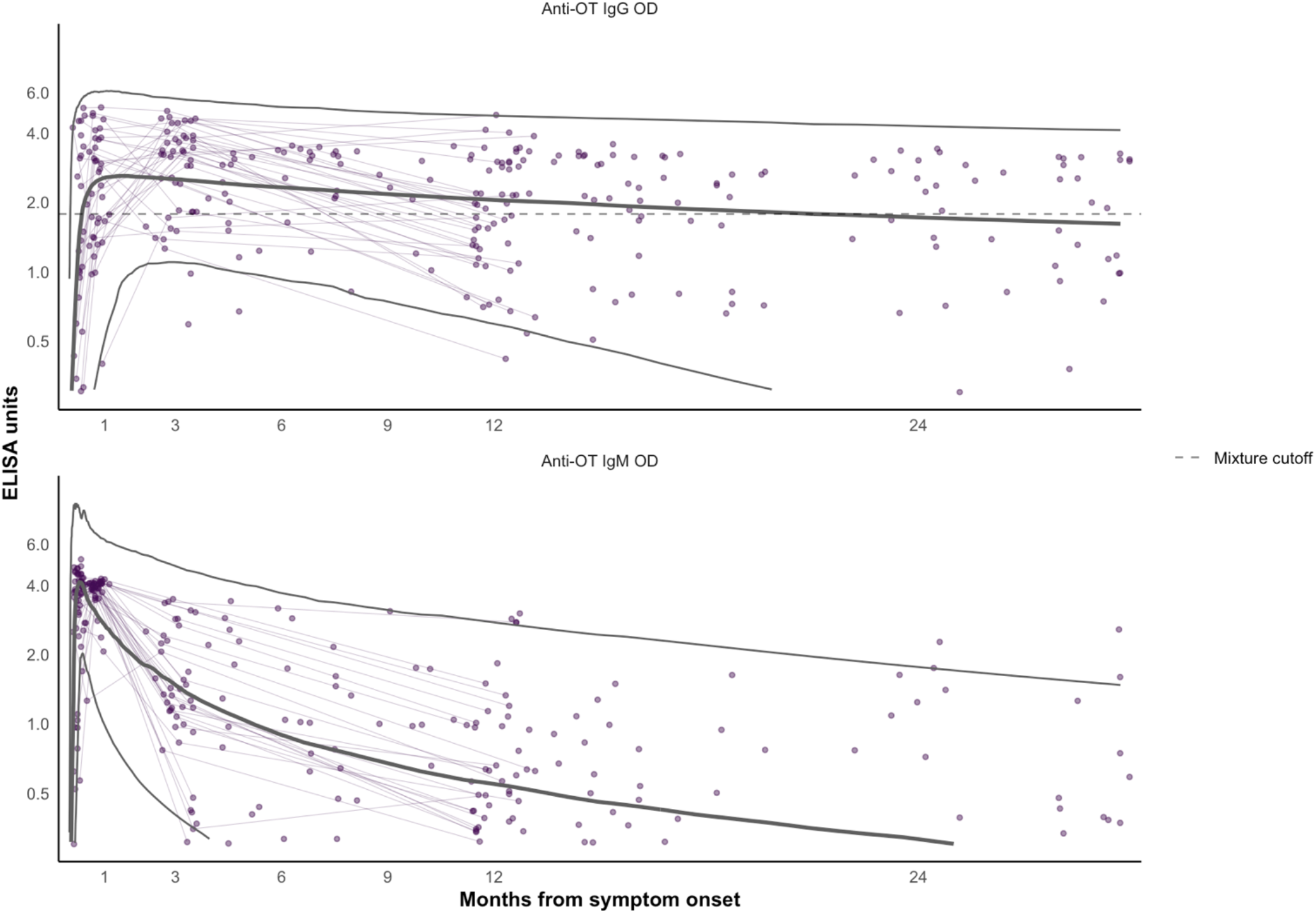
Kinetics of IgM responses among Scrub Typhus cases. Longitudinal antibody dynamics were modeled from ELISA-measured antibody responses using Bayesian hierarchical models. The points are the observed individual antibody concentrations; each point indicates one sample; a patient’s samples are connected with light purple lines. The dark solid lines indicates the median, and the 95% credible intervals for the model-fitted antibody decay concentrations. The horizontal dashed line denotes the mixture model cutoff.

For the population data, ELISA antibody responses were measured from 721 participants in Vellore, India, and 1105 participants in Kathmandu and Kavre districts, Nepal. The median age of participants was 49 in India (IQR 40-62) and 11 in Nepal (IQR 5.5-17). In India, 63.2% (456/721) of participants were female, compared to 48.7% (538/1105) in Nepal (Table 2).

**Table 2:**
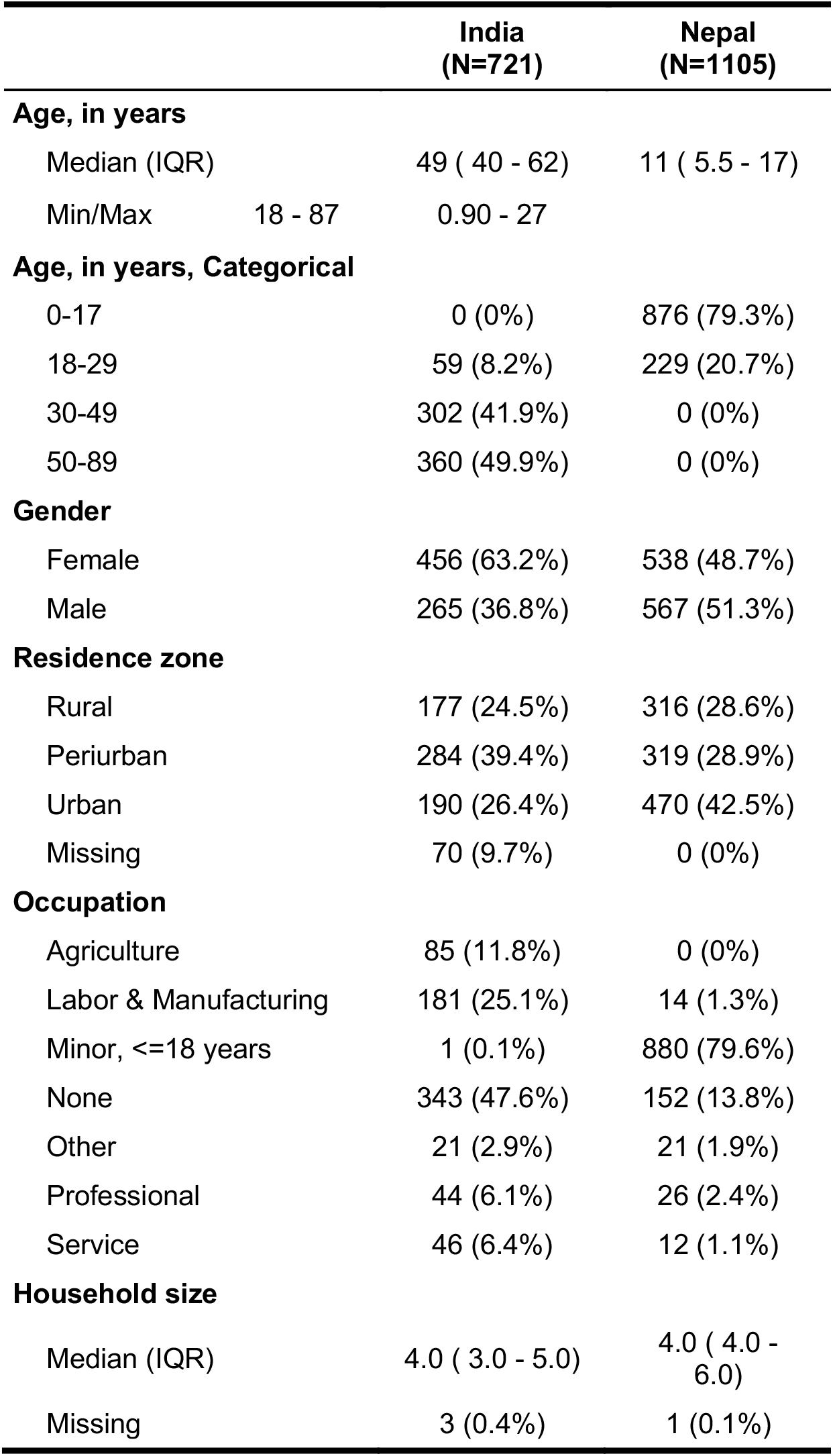
Population data summary.

|  | India<br>(N=721) | Nepal<br>(N=1105) |
| --- | --- | --- |
| **Age, in years** |  |  |
| Median (IQR) | 49 ( 40 - 62) | 11 ( 5.5 - 17) |
| Min/Max 18 - 87 | 0.90 - 27 |  |
| **Age, in years, Categorical** |  |  |
| 0-17 | 0 (0%) | 876 (79.3%) |
| 18-29 | 59 (8.2%) | 229 (20.7%) |
| 30-49 | 302 (41.9%) | 0 (0%) |
| 50-89 | 360 (49.9%) | 0 (0%) |
| **Gender** |  |  |
| Female | 456 (63.2%) | 538 (48.7%) |
| Male | 265 (36.8%) | 567 (51.3%) |
| **Residence zone** |  |  |
| Rural | 177 (24.5%) | 316 (28.6%) |
| Periurban | 284 (39.4%) | 319 (28.9%) |
| Urban | 190 (26.4%) | 470 (42.5%) |
| Missing | 70 (9.7%) | 0 (0%) |
| **Occupation** |  |  |
| Agriculture | 85 (11.8%) | 0 (0%) |
| Labor & Manufacturing | 181 (25.1%) | 14 (1.3%) |
| Minor, <=18 years | 1 (0.1%) | 880 (79.6%) |
| None | 343 (47.6%) | 152 (13.8%) |
| Other | 21 (2.9%) | 21 (1.9%) |
| Professional | 44 (6.1%) | 26 (2.4%) |
| Service | 46 (6.4%) | 12 (1.1%) |
| **Household size** |  |  |
| Median (IQR) | 4.0 ( 3.0 - 5.0) | 4.0 ( 4.0 - 6.0) |
| Missing | 3 (0.4%) | 1 (0.1%) |

In both India and Nepal, the seroprevalence of scrub typhus infection increased with age (Figure 3). In Vellore, India the overall seroprevalence was 29.8% (95% CI 26.5-33.2): growing from 10.2% (95% CI 2.4-17.9) among 18-29-year-olds to 38.1% (95% CI 33.0-43.1) among 50 to 89-year-olds. In Kathmandu and Kavre, Nepal, the overall seroprevalence was 1.2% (95% CI 0.5-1.8), rising from 0.1% (95%CI 0-0.3) among 0 to 17-year-olds to 5.2% (95%CI 2.3 – 8.1%) among 18-29-year-olds (Table 3).

**Table 3:** Age-specific seroprevalence and seroincidence in Nepal and India.

| Seroincidence per 1000 person-years (95% CIs) |  |  |  |  |  |  |
| --- | --- | --- | --- | --- | --- | --- |
| Age | N | N IgG seropositive | IgG seroprevalence (95% CI) | IgG antibody kinetics | IgG and IgM antibody kinetics | Derivative of age-dependent IgG seroprevalence |
| India |  |  |  |  |  |  |
| 0-17 | 0 | 0 | – | – | – | – |
| 18-29 | 59 | 6 | 10.2% (2.4-17.9) | 8 (4-16) | 11 (6-18) | 4 (2-10) |
| 30-49 | 302 | 72 | 23.8% (19.0-28.7) | 14 (11-18) | 12 (10-15) | 7 (5-9) |
| 50-89 | 360 | 137 | 38.1% (33.0-43.1) | 26 (21-32) | 20 (17-24) | 8 (6-9) |
| Overall | 721 | 215 | 29.8% (26.5-33.2) | 18 (16-21) | 16 (14-18) | 7 (6-8) |
| Nepal |  |  |  |  |  |  |
| 0-17 | 876 | 1 | 0.1% (0-0.3) | 2 (1-4) | – | 0 (0-1) |
| 18-29 | 229 | 12 | 5.2% (2.3-8.1) | 9 (6-14) | – | 3 (1-4) |
| 30-49 | 0 | 0 | – | – | – | – |
| 50-89 | 0 | 0 | – | – | – | – |
| Overall | 1105 | 13 | 1.2% (0.5-1.8) | 4 (3-6) | – | 1 (1-2) |

**Figure 3:**
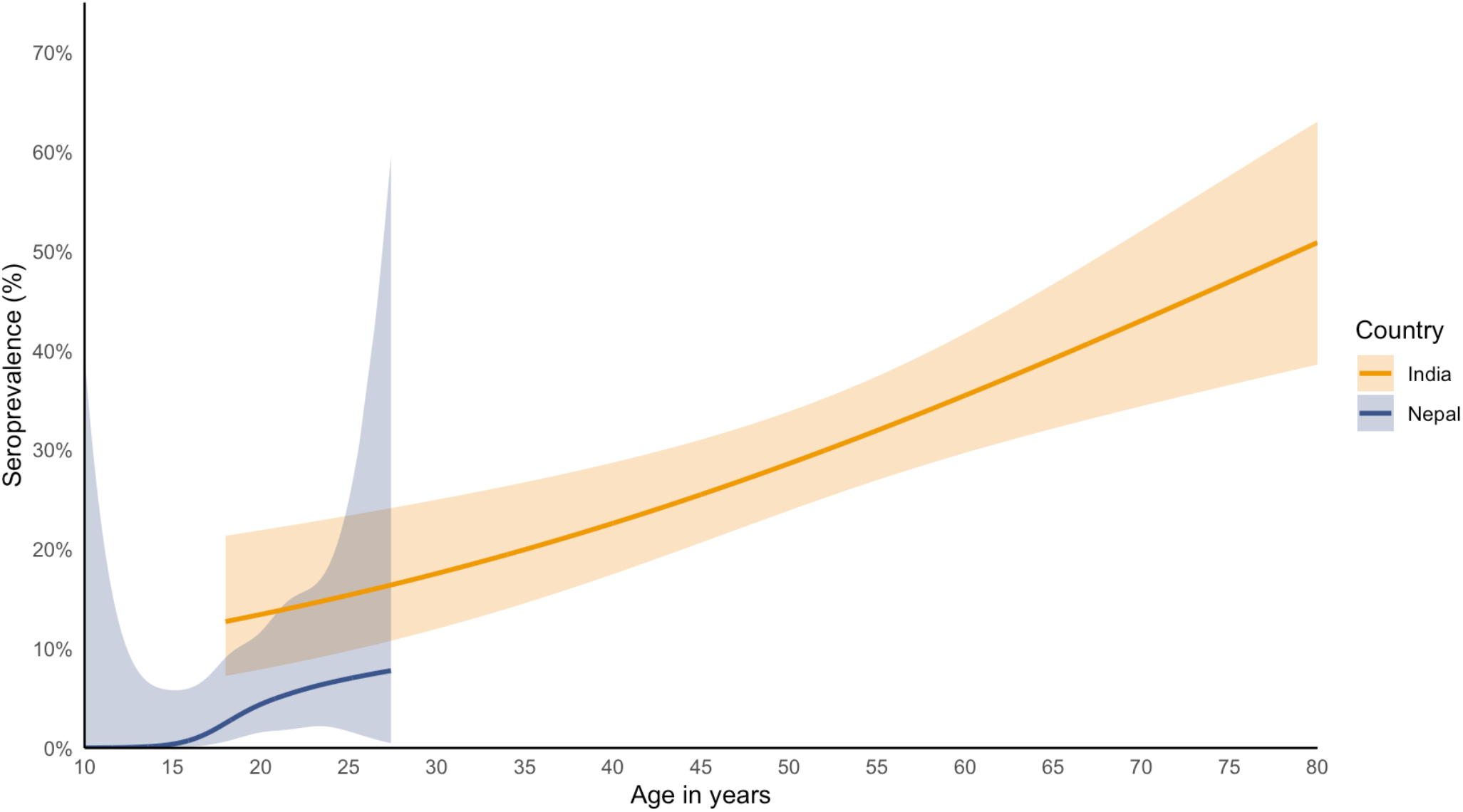
Age-dependent seroprevalence. Seroprevalence is depicted as a function of age. Seroprevalence was determined using the mixture-model derived cutoff for IgG responses. The age-dependent seroprevalence was modeled using a generalized additive model with a cubic spline for age and simultaneous confidence intervals using a parametric bootstrap of the variance-covariance matrix of the fitted model parameters.

In Vellore, India, the overall scrub typhus seroincidence rate was 18 per 1000 person-years (95% CI: 16 - 21) using IgG antibody dynamics. Seroincidence rose with age; growing from 8 (95% CI 4 - 16) among 18-29 year-olds to 26 (95% CI 21 - 32) among 50-89-year-olds. In the Kathmandu valley of Nepal, the overall seroincidence was 4 (95% CI 3 – 6) per 1000 person-years, also rising with age from 2 (95% CI 1 - 4) per 1000 person-years among 0 to 17-year-olds to 9 (95%CI 6 -14) among 18 to 29-year-olds. In both Nepal and India, the overall seroincidence using antibody dynamics was more than double the seroincidence derived from the age-dependent IgG-seroprevalence, 2.6 times higher in India and 4 times higher in Nepal (Table 3). IgM responses were only available from the study in India; there, the seroincidence using IgM was similar to IgG. The confidence intervals using IgM and IgG together were more narrow than using IgG alone.

In both sites, scrub typhus seroincidence varied by resident zones. In Vellore, India, seroincidence was highest in periurban settings across all age groups, followed by rural and then urban, whereas in Nepal, seroincidence was higher in urban settings (Figure 4). In India, females had slightly elevated seroincidence rates compared to males across all age groups, although the confidence intervals overlapped (Figure 4).

**Figure 4:**
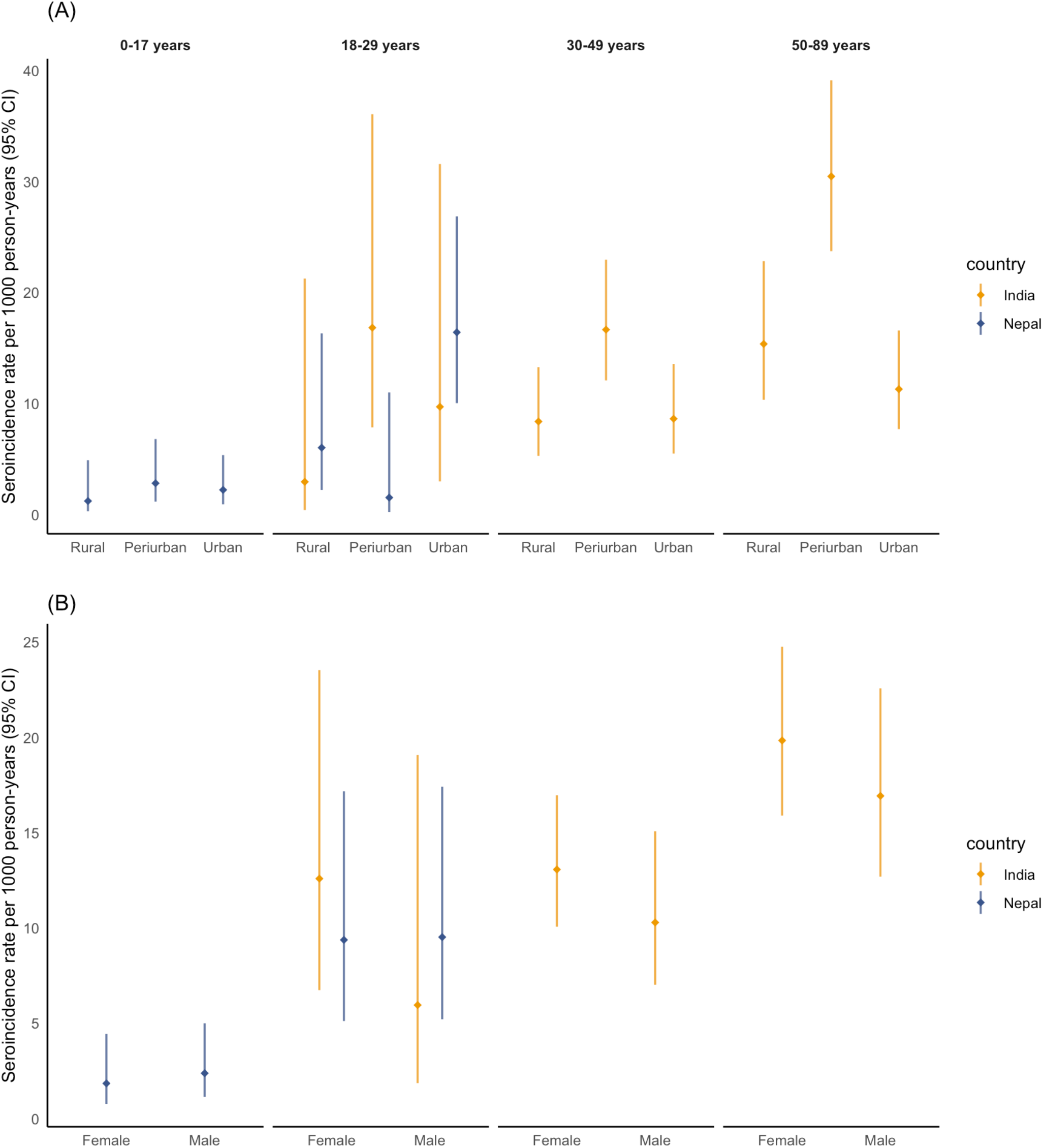
Seroincidence by age plus (A) resident zones and (B) gender. Seroincidence rates across are shown across age strata, residence zone (A), and gender (B) comparing the India and Nepal population samples. Seroincidence is estimated using IgG antibody kinetics. The point represents the incidence estimate per 1000 person-years, and the bar indicates the 95% confidence interval.

## Discussion

In this study, we developed and applied a novel analytic approach to estimate the seroincidence of scrub typhus using antibody decay information from confirmed cases in Thailand and India and cross-sectional population serosurveys in Nepal and India. Seroincidence, the number of new infections in a population per year, characterizes transmission intensity and is highly informative for determining where and among whom infection burden is highest. We found a high seroincidence of scrub typhus in both study populations, underscoring the importance of addressing the burden of scrub typhus infection in these regions. A key finding of our study involves delineating the persistence of antibodies after infection, characterizing their decay rate and shape, and using this information to interpret population-level responses. Our results demonstrate that employing the cutoff method to estimate seroprevalence while disregarding antibody decay underestimates the true burden of scrub typhus infection.

Most cross-sectional seroepidemiologic studies on scrub typhus rely on seroprevalence as a marker of infection, employing a cutoff threshold to dichotomize IgG responses and calculating the proportion of the population with values above the threshold (6,8,31). Antibody responses in a population at any given time will be a function of exposure, the age of the population, and antibody persistence. For example, in our study populations, the overall seroprevalence was much higher in India than in Nepal; however, in India, the median age of patients was 49 years compared to the median age of 11 years in Nepal. Despite differences in overall seroprevalence rates, the discrepancies in age-specific seroprevalence rates between the two populations were less marked. This underlines the importance of considering age-specific seroprevalence rates when comparing seroprevalence between different populations, as the age distribution can have a considerable impact on the observed seroprevalence. Our novel analytical approach, which accounts for age and antibody decay, provides a more accurate representation of scrub typhus infection in these regions, highlighting the need for targeted public health interventions and research.

In both India and Nepal, seroprevalence increased notably with age, a finding that has been consistently observed in other scrub typhus serosurveys (17,32). An increase in seroprevalence with age can result from gradual accumulating in IgG responses over time or from a genuinely elevated risk of infection in older age groups. Both arguments have been put forth for scrub typhus seroepidemiology, with the suggestion that elevated risk in older ages is attributable to changes in skin health, immune system function, and physiological processes (29). We argue that reporting seroincidence rather than seroprevalence is essential to disentangling the effects of IgG accumulation over age versus age-specific risk. Seroincidence, which measures the number of new infections in a population per year, characterizes transmission intensity and is highly informative for determining where and at what ages the infection burden is highest. Incorporating antibody dynamics into the calculation of seroincidence, as demonstrated in this paper, further elucidates age-specific changes in infection risk by accounting for decaying antibody levels. In our study populations, we observed a marked increase in seroprevalence with age, while the increase in seroincidence at older ages was comparatively more gradual. For example, in India, individuals aged 50-89 had 3.8 times higher IgG seroprevalence compared to 18-29 year-olds compared to 1.8 times higher seroincidence using IgG and IgM antibody dynamics. This highlights the importance of using seroincidence to better understand the age-specific risk of infection and to inform targeted public health interventions.

Defining antibody dynamics and persistence is crucial for interpreting population-level seroresponses to Orientia tsutsugamushi and modeling seroincidence. In our study, we found that median IgG responses remained elevated above a mixture-model cutoff of 1.7 OD for nearly two years. These findings align with a recent study by Schmidt et al., which reported that half of the cases had persistent IgG antibodies beyond two years (29). Interestingly, IgM responses persisted longer than anticipated. We observed that half of the participants had an OD <1 after 5.3 months after symptom onset, and 90% of participants had an OD <1 for 26.5 months. This extended persistence contrasts with Schmidt et al’s findings, where half of the participants had IgM OD <1 after 2.7 months and 90% had OD <1 after 7.7 months. A potential explanation for this discrepancy could be the difference in the duration of sample collection between the two studies. In the Schmidt study, samples were collected up to a year after infection, while our study included samples collected up to 3.5 years after infection. Another explanation could be the differences in modeling approaches. Our power-function model allows for the shape of the decay to be non-exponential, permitting a slower decay at longer times since infection. In contrast, a restricted cubic spline approach may require a constant rate and shape of decay. It would be ideal to test these approaches with the same data in future research. The persistence of IgM revealed in our study suggests that it would be a good complement to IgG to identify recent infections and improve the precision around seroincidence estimates. Indeed, for the India data where IgM results were available, the seroincidence estimates using both IgM and IgG were consistent, yielding similar estimates for the seroincidence, thus strengthening the validity of our approach. The seroincidence estimates using IgM and IgG had narrower confidence intervals than those using IgG alone.

One of the key strengths of our approach lies in its ability to account for the inherent heterogeneity of antibody responses, as well as measurement and biologic noise. Traditional seroepidemiological methods, which rely on a single cutoff value for seroprevalence and seroincidence, fail to incorporate these sources of variability. By explicitly integrating the heterogeneity in antibody responses, our method captures the distribution of responses among individuals across ages, clinical severity and levels of immune function. Additionally, our approach takes into account the measurement noise associated with the ELISA assay and the biologic noise that stems from non-specific antibody binding. This more comprehensive incorporation of uncertainty leads to more accurate and robust estimates of scrub typhus seroincidence, which in turn better informs public health interventions and research priorities.

Our findings have several implications for public health. First, we reveal a substantial subclinical burden of scrub typhus beyond what is captured by the reporting of clinical cases. In India, a recent population-based fever surveillance study conducted reported a clinical incidence of 0.8 cases per 1000 person-years, defined as the incidence of scrub typhus among febrile individuals who sought care or were hospitalized (32). By applying our seroincidence estimates derived from IgG and IgM antibody dynamics, we estimate that there are around 14 subclinical infections for every clinical case in the population surrounding CMC Hospital. This inflation factor is less than estimated by Devamani et al., who reported a seroincidence of 44 per 1000 person-years. This study was conducted in a population that was purposefully selected for its elevated numbers of clinical scrub typhus cases, which could explain the differences (30). Population-based clinical incidence estimates for scrub typhus are not currently available for the study population in Nepal but would be valuable in distinguishing clinical versus sub-clinical infections. While individual health implications of subclinical infections are currently unclear, seroincidence studies like ours are useful for identifying areas with substantial scrub typhus burden, thereby informing the allocation of resources for enhanced clinical diagnostic capacity and targeted surveillance efforts. Furthermore, our study challenges the notion that scrub typhus is solely a rural disease. We observed elevated incidence rates in both urban and peri-urban communities in Nepal and India. However, we could not ascertain whether exposure occurred at the place of residence or was linked to rural exposure, highlighting the need for further research targeting these populations and investigating exposures. Expanding our understanding of the distribution and risk factors associated with scrub typhus in various settings will help inform the development of more effective and targeted prevention and control measures, ultimately reducing the burden of disease in affected populations.

There are several limitations to our study that warrant consideration. First, the population serosurveys in India and Nepal were conducted over different time periods, with the Indian survey spanning a few months and the Nepalese survey taking place over a year. This discrepancy could introduce variability in the observed seroprevalence and seroincidence rates between the two populations due to seasonal differences in scrub transmission. Second, our study assumes that the antibody decay patterns observed in the cohorts from Thailand and India are applicable to the populations in Nepal and India. This may not necessarily hold true, as factors such as age, geography, clinical severity, and population-specific immune characteristics could lead to variations in antibody decay patterns. Third, we estimated antibody dynamics using cases from a wide range of ages. Although our sample size was not large enough to investigate age-specific variability in kinetics, it is possible that these dynamics may vary with immune function at different ages, presenting an interesting avenue for future research. Additionally, our findings may be influenced by the overrepresentation of higher-risk groups in our data, such as women and urban residents, which could affect the population-level seroincidence estimates. Future studies may benefit from adjusting for potential biases introduced by overrepresented demographic groups. Lastly, we do not have sufficient data to investigate whether antibody decay rates vary by age, which could influence our seroincidence estimates. Despite these limitations, our study offers valuable insights into the seroepidemiology of scrub typhus and highlights the importance of accounting for antibody dynamics and heterogeneity in such analyses.

Scrub typhus remains an important but underrecognized etiology of acute fever with endemicity expanding globally. There is a critical need for low-cost, accurate tools to quantify the burden of scrub typhus infections to inform public health decision-making. We describe a sero-surveillance approach that can efficiently generate population-level scrub typhus seroincidence estimates from cross-sectional serosurveys. This methodology offers a valuable new tool for informing targeted prevention and control strategies, ultimately contributing to a more effective response to scrub typhus in endemic regions worldwide.

## Data Availability

Before publication we will make all de-identified data available via Open Science Framework.

## Acknowledgments

We gratefully acknowledge the study participants for their valuable time and interest in participating in these studies.

## Supplemental material

**Supplemental Figure 1:**
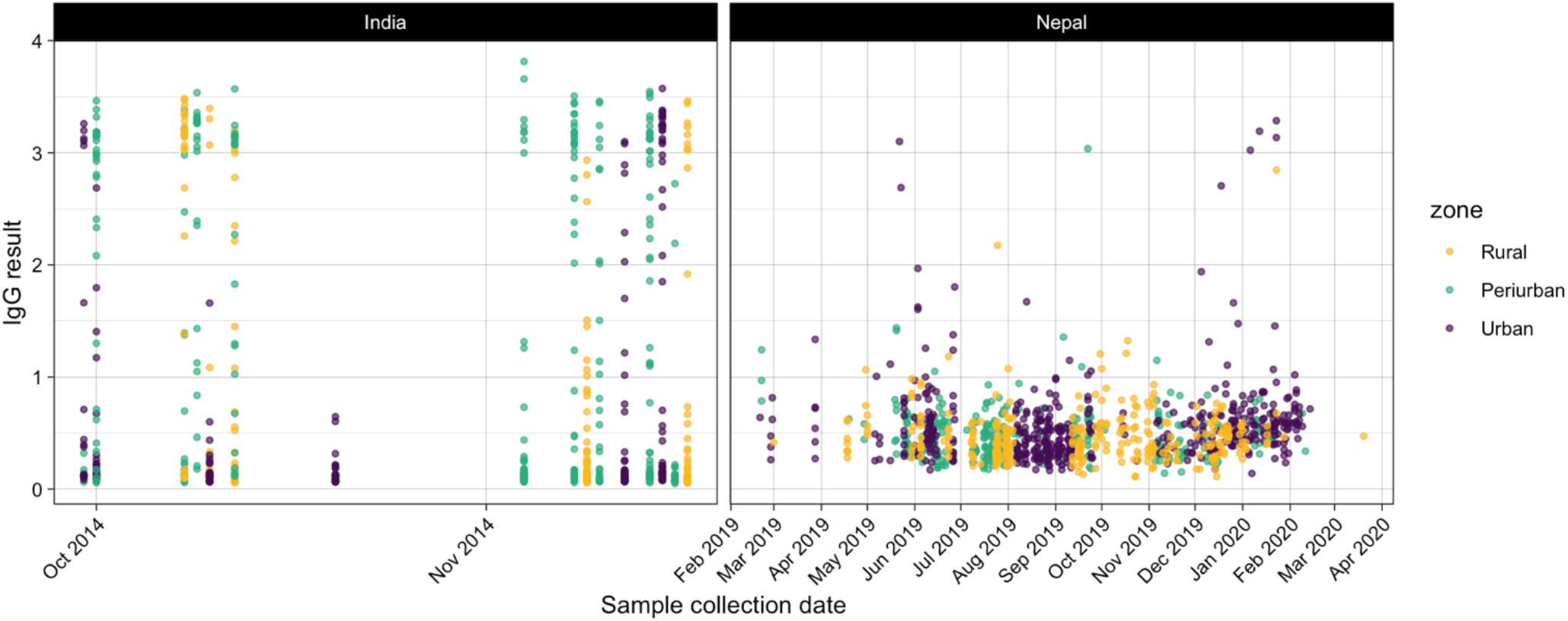
Sampling collection over time and geographic unit

**Supplemental Table 1:**
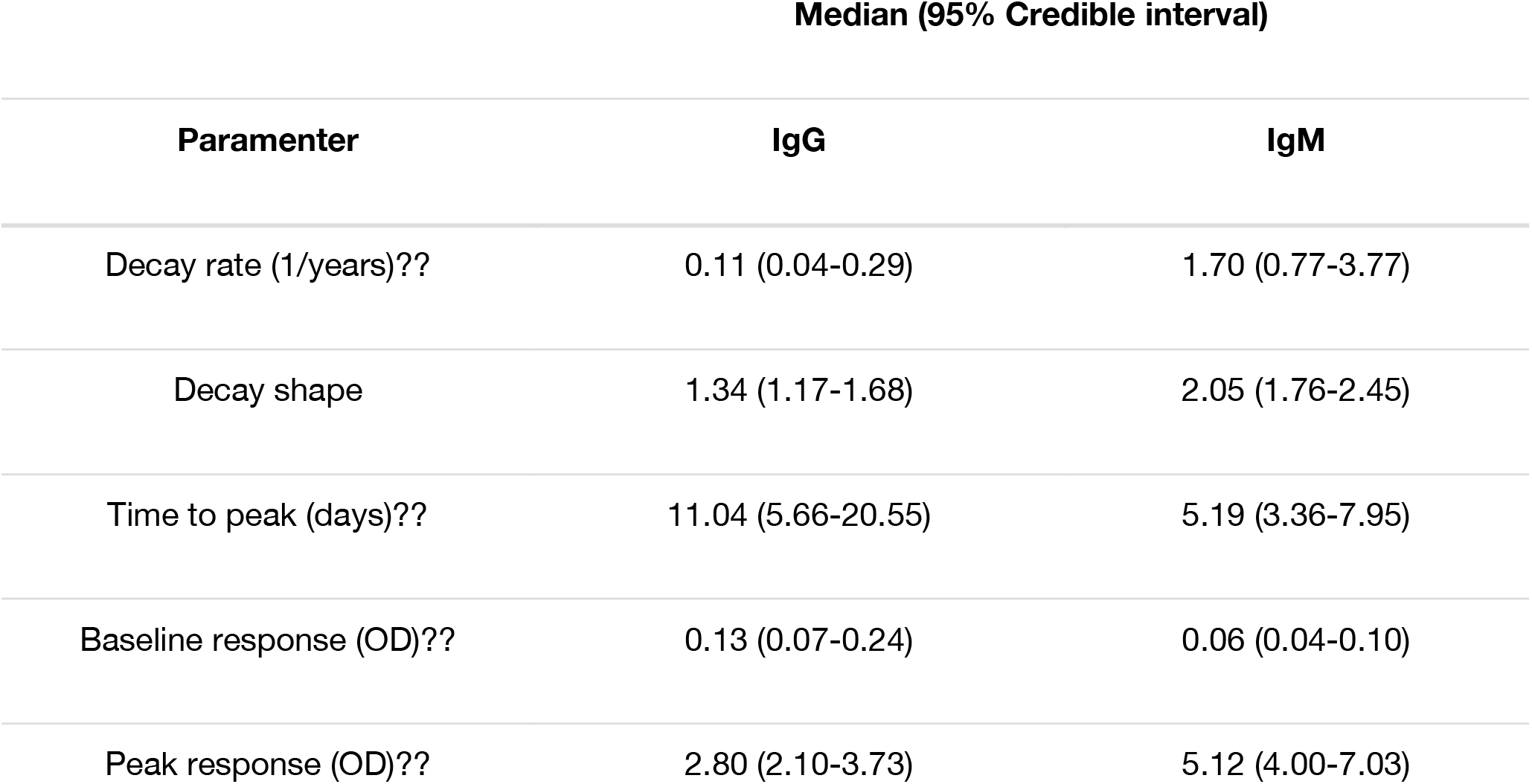
Modeled antibody kinetic parameters.

